# Application of syringe-based device for pressure pain threshold measurements under autoalgometry settings

**DOI:** 10.64898/2026.10.13.26365379

**Authors:** S. Frankevich, I. Michaelevski, D. Yakubovich

## Abstract

**Background:** Pressure pain threshold (PPT) is commonly assessed using pressure algometry. Self-administered algometry could reduce examiner involvement and may facilitate repeated assessment outside specialist settings.

**Objective:** To examine the concurrent agreement of a previously described syringe-based pneumatic algometer with a commercially available comparator during self-administered PPT measurement in healthy adults.

**Methods:** Thirty healthy volunteers completed researcher-administered and self-administered pain threshold measurements utilizing an AMF-500 device and the syringe-based pneumatic algometer.

**Results:** During self-administration, measurements from the two devices were strongly correlated (r = 0.82, p < 0.05); the mean paired difference was 6.01 N (95% CI, -0.58 to 12.59 N). Self-administered test values were lower than researcher-administered test values in the fixed-order comparison (p = 0.0029).

**Conclusion:** According to our findings the proposed device is a valid tool for autoalgometry and may potentially be introduced as an element of routine self-assessment of PPT.

## Introduction

Pain is a complex sensation involving nociceptive, cognitive, and affective processes (1). It is a common clinical symptom, reported to be the most frequent cause of admission to primary care (1), and is a useful diagnostic criterion. The experience of pain is inherently subjective and cannot be measured directly; instead, clinicians and researchers lean on the patient’s own perception. Several methods of pain evaluation are currently in use (2). One such method relies on measuring the intensity of an external stimulus that is perceived as pain, known as pain threshold (3).

One of the most widely used methods of pain threshold evaluation is Fischer pressure algometry (4). An examiner presses an algometer, against the skin of the subject in a predetermined spot. The point at which the increasing pressure transforms into a subjective feeling of pain is recorded as pain-pressure threshold (PPT). Pressure algometers have been validated for use in the clinic and in research (5-8), and models based on several methods of pressure or applied force evaluations have been developed. To increase accuracy and reproducibility of PPT measurement, a method of autoalgometry has been proposed (9, 10).

Autoalgometry devices are designed to allow the subject to perform PPT evaluation without assistance from examiner. This is achieved by affixing the pressure probe to a stationary frame, with the subject pressing against the probe, usually at the base or phalanx of a finger. Resulting pressure is recorded digitally. During standard pressure threshold measurement time from the subjective feeling of pain to cessation of the test includes reaction time of the examiner. In contrast autoalgometry is independent of examiner response time thus resulting in PPT value which is closer to the “true” pain threshold. In addition, because autoalgometry does not involve a medical professional, it may be adopted as a method of routine evaluation of PPT in patients for whom pain perception is diagnostically valuable, such as in case of peripheral neuropathy.

Peripheral neuropathy is associated with numerous conditions, including diabetes mellitus, hypothyroidism, inflammatory disorders, nutritional deficiencies, and chemotherapy-induced neurotoxicity (11). Early discovery and constant monitoring of neuropathic dysfunction, including altered pain perception, are essential for treatment and relief of peripheral neuropathy (12, 13). There is, however, no established practice of at-home self-evaluation of pain perception for patients with peripheral neuropathy. Autoalgometry may fill this gap as a safe, accurate and easy to perform method of PPT evaluation. In this work we validate a previously developed (14) algometry device based on a pneumatic cylinder for use in autoalgometry against a commercial algometer taken as a reference comparator. No patient cohort, longitudinal assessment, or home-use evaluation was included in the present study.

## Materials and Methods

An a-priori sample size estimation was conducted to determine the minimum number of study participants required to accurately evaluate concurrent validity between the custom algometry device and the common comparator (the AMF-500) during self-administration. We conducted power analysis based on Cohen’s Standard Effect Size (15). The sample size (N) is calculated according to Eq.1

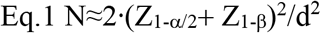

where Z_1-α/2_− standard normal deviation for certain significance, at α = 0.05 this value is 1.96, Z_1-β_ is standard normal deviation for desired statistical power. For 80% power (β= 0.20), this value is 0.84, *d* is Cohen’s effect size (for large effect size was taken 0.8). The above-described calculations rendered *N* ≈25.

A total of 30 healthy adult participants (14 male, mean age 29.4±0.7; 16 female, mean age 33.8±2.3) were recruited from among students and staff of Ariel University on a volunteer basis. All participants received and signed an informed consent form which detailed study aims and experimental procedures. Exclusion criteria were currently experiencing acute or chronic pain and currently taking analgetic medication. The study was approved by the Ariel University Medical School Ethical Committee (approval number AU-MED-DY-20231219).

The pneumatic algometry device described in this study is a syringe based algometer we developed previously (14). The custom device consists of a 60-mL Luer-lock plastic syringe (BD Plastipak, USA) **(Fig. 1A)**, a 1-cm^2^ square probe fixed to the plunger thumb rest, and an NUL-210 16-bit 100 Hz pressure sensor (NeuLog, Israel). Internal air pressure was streamed by USB to a personal computer for analysis. For comparison with the AMF-500, the device output was converted from internal pressure (kPa) to applied force (N) using the previously published calibration equation (14). The AMF-500 was equipped with a pressure probe of identical construction and area.

**Figure 1.**
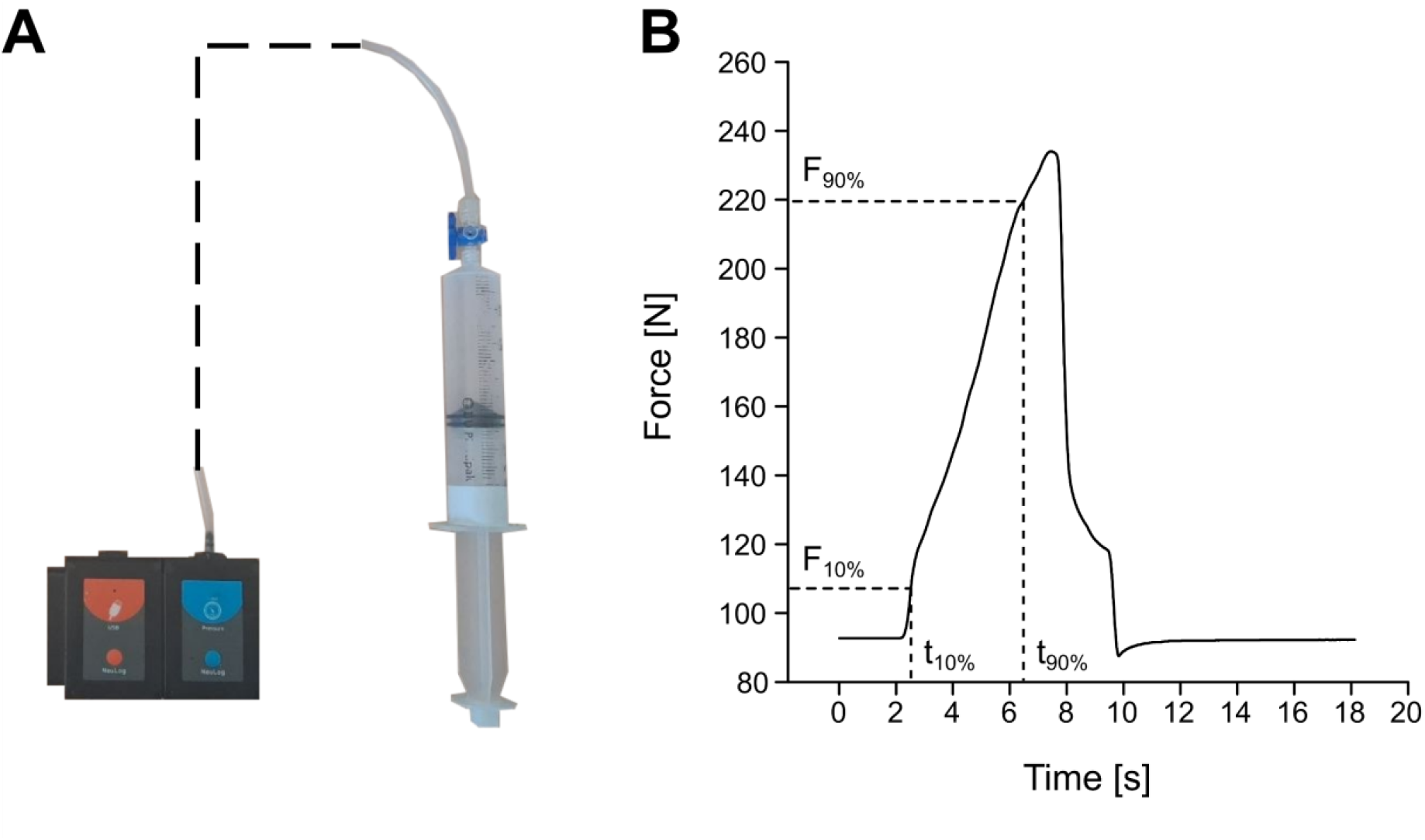
The pneumatic autoalgometry device. **A.** Representative image of the device. The air-tight plastic syringe is connected to the NUL-210 100 Hz pressure sensor through a flexible plastic tube (dashed line). **B**. Representative plot of applied force time-course. Dotted lines represent points corresponding to 10% (F_10%_) and 90 % (F_90%_) of maximal response (after baseline subtraction). t_10%_ and t_90%_ are correspondently the time points at which the above-mentioned pressures were measured.

We evaluated concurrent validity of our device with an existing algometry device (AMF-500, Beslands, China). Assessment of pain-pressure threshold (PPT) was performed in a predetermined spot at the midpoint of the forearm over the radius. Four tests were conducted in the following order: AMF-500, applied by the researcher; AMF-500, applied by the volunteer; custom algometry device, applied by the researcher; custom algometry device, applied by the volunteer. We chose a resting period of 60s between applications (16, 17).

PPT evaluation was performed along commonly practiced guidelines (4). The participants were first introduced to the AMF-500 and the custom algometry device and allowed to practice force application on an inert flat surface. The participants were asked to sit in a chair and position their dominant arm in front of them resting on a table with the radial side facing upward. Using one hand to apply pressure and another stabilizing the device, the researcher pressed the probe against the algometry point with the device oriented perpendicularly to the surface of the skin. Once the volunteer reported the first feeling of pain, the procedure was stopped immediately. The self-administration test was carried out in the same manner, with the volunteer using their non-dominant arm to apply pressure to their dominant arm. The maximum logged force (AMF-500) and pressure (custom algometry device) were interpreted as PPT. For this purpose, the AMF-500 was used in a mode which registered the highest applied force before manual reset. All data were streamed to a personal computer through a USB connection utilizing official NeuLog software (NeuLog data acquisition and analysis application v. 7.323.153).

To evaluate concurrent validity and potential application bias of the custom algometry device relative to the reference comparator (AMF-500), a multi-step analysis was performed. All descriptive statistics (Mean, SD, Median) were computed for data obtained. Concurrent validity between devices during self-administered tests (autoalgometry) was assessed using the Pearson correlation coefficient (*r*) and Bland-Altman plot analysis to evaluate mean bias and 95% limits of agreement (±1.96 SD).

Temporal force dynamics were evaluated by calculating the 10-90% rise time (t_10-90_) and rate of change of force 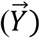(**Fig. 1B**) were calculated according to Eq.2 and Eq.3

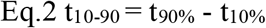

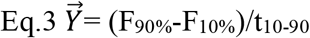

where t_90%_ and t_10%_ are corresponding time points at which force of F_90%_ and F_10%_ relative to PPT were measured.

Since same persons underwent several procedures during testing, we conducted repeated measures ANOVA analysis in order to estimate the contribution of several parameters (device type, operator (researcher/self), gender of the volunteer) to PPT, t_10-90_ and 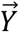.

Statistical analysis was performed utilizing Sigmaplot 16 (Grafiti LLC, USA) and Jamovi (Edelsbrunner et al., Open Source). Outliers (1 measurement) was detected using Tukey fences method (18, 19). Another measurement was removed due to incomplete data availability (t_10-90_ was absent). After outliers and missing data removement 28 volunteers’ data was subjected to complete analysis.

## Results

In order to estimate validity of utilization our device and to examine possible bias we conducted correlation analysis and constructed Bland-Altman plots. Pain-pressure thresholds (PPT) obtained via custom device and reference comparator (AMF-500) demonstrated strong concurrent validity with Pearson’s correlation >0.8 both in self-administered test and researcher administered test **(Fig. 2A** and **2B)**. According to Bland-Altman plot (**Fig. 2C** and **2D**) it can be see that no substantial bias was demonstrated both in measurements conducted by researcher and self-obtained values.

**Figure 2.**
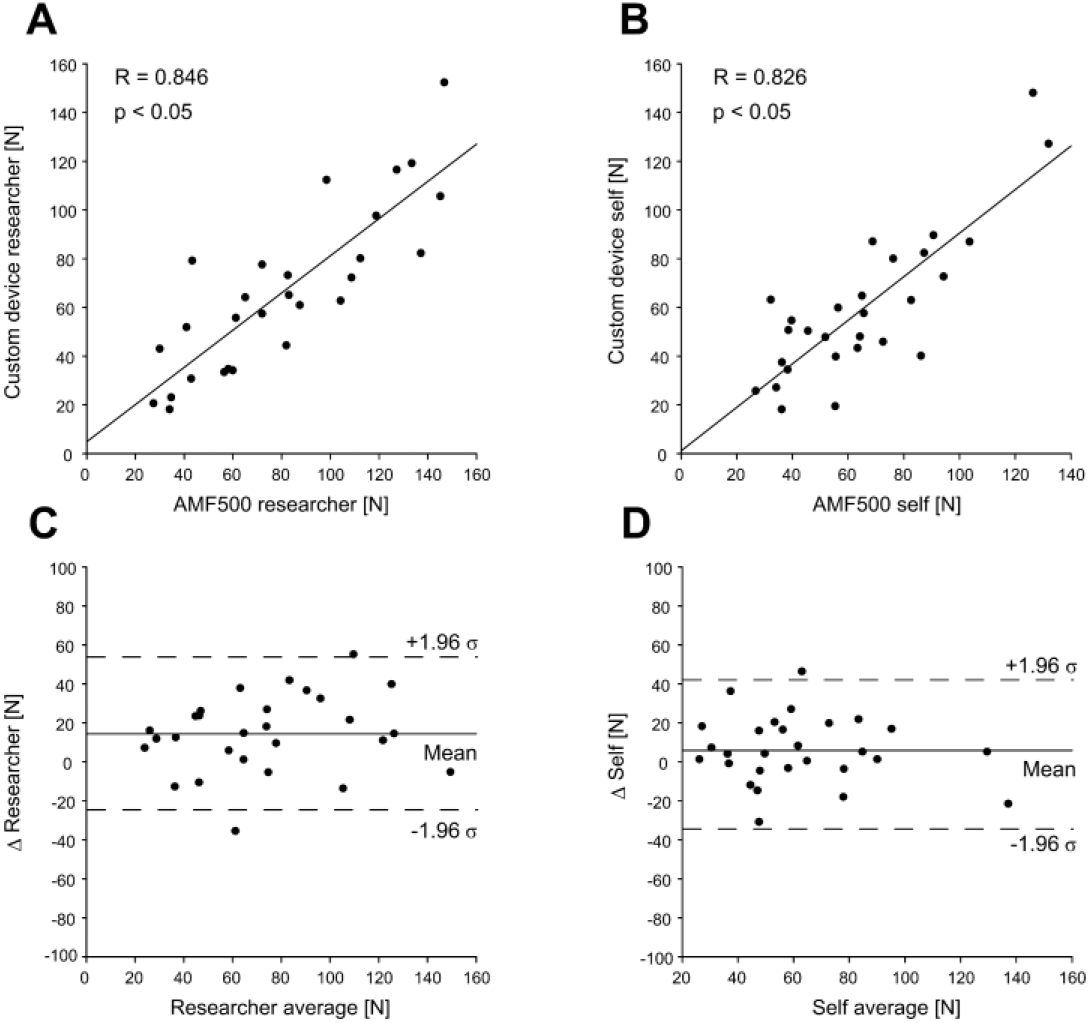
Validation of the pneumatic algometry device against a commercially available pressure algometer. **A.** Correlation analysis of pain-pressure threshold data (PPT) obtained by researcher (he AMF-500 and the custom algometry device). **B**. Correlation analysis of pain-pressure threshold data (PPT) obtained by volunteers (self, AMF-500 and the custom algometry device). **C**. Bland– Altman plot of researcher-administered measurements, displaying mean paired bias and 95% limits of agreement (mean difference ± 1.96σ). **D**. Bland–Altman plot of self-administered measurements, displaying mean paired bias and 95% limits of agreement (mean difference ± 1.96σ).

We evaluated possible factors which may affect PPT measurements **(Tab. 1, Fig.3A)**. As described above (Materials and Methods) we conducted repeated measures ANOVA analysis defining the within subject effects as device type (AMF-500 versus custom algometer), method of measurement (researcher versus self) and between subject effect as gender.

Based on data in Table 1 both effects of device and method were significant while the effect of gender was not significant.

**Table 1.** Results of the repeated measures ANOVA analysis evaluating effects of device choice and method choice on pressure-pain threshold (PPT) measurements.

| Within subject effects | Sum of Squares | df | Mean Square | F | p |
| --- | --- | --- | --- | --- | --- |
| Device | 3030 | 1 | 3030 | 20.21 | **< 0.001** |
| Device vs Gender | 274 | 1 | 274 | 1.83 | 0.188 |
| Residual | 3899 | 26 | 150 |  |  |
| Method | 3234 | 1 | 3234 | 9.51 | **0.005** |
| Method x Gender | 536 | 1 | 536 | 1.58 | 0.220 |
| Residual | 8841 | 26 | 340 |  |  |
| Device x Method | 598 | 1 | 598 | 3.16 | 0.087 |
| Device x Method vs Gender | 248 | 1 | 248 | 1.31 | 0.263 |
| Residual | 4925 | 26 | 189 |  |  |
| Between subject effects | Sum of Squares | df | Mean Square | F | p |
| Gender | 2848 | 1 | 2848 | 0.814 | 0.375 |
| Residual | 90999 | 26 | 3500 |  |  |

To further explore differences resulting from the application technique, we analyzed the temporal dynamics of internal air pressure rise in the pneumatic algometry device by calculating the 10-90% rise time for the custom algometry device. Additionally, we calculated loading rates for each method of application. Loading rate was defined as (F_90%_-F_10%_)/(t_90%_- t_10%_) (**Fig. 1B**). The rise-time values showed no significant differences between researcher-administered tests and autoalgometry (**Tab. 2**); the loading rates, however, were significantly dependent on measurement method (**Tab. 3**).

**Table 2.** Results of the repeated measures ANOVA analysis evaluating effects of method choice and volunteer gender on t_10-90_.

| Within subject effects | Sum of Squares | df | Mean Square | F | p |
| --- | --- | --- | --- | --- | --- |
| Method | 13.950 | 1 | 13.950 | 3.365 | 0.078 |
| Method vs Gender | 0.575 | 1 | 0.575 | 0.139 | 0.713 |
| Residual | 107.782 | 26 | 4.145 |  |  |
| Between subject effects | Sum of Squares | df | Mean Square | F | p |
| Gender | 17.8 | 1 | 17.8 | 1.44 | 0.240 |
| Residual | 320.4 | 26 | 12.3 |  |  |

**Table 3.** Results of the repeated measures ANOVA analysis evaluating effects of method choice and volunteer gender on loading rate.

| Within subject effects | Sum of Squares | df | Mean Square | F | p |
| --- | --- | --- | --- | --- | --- |
| Method | 266.5 | 1 | 266.5 | 5.911 | **0.022** |
| Method vs Gender | 24.1 | 1 | 24.1 | 0.534 | 0.471 |
| Residual | 1172.4 | 26 | 45.1 |  |  |
| Between subject effects | Sum of Squares | df | Mean Square | F | p |
| Gender | 430 | 1 | 430 | 0.795 | 0.381 |
| Residual | 14069 | 26 | 541 |  |  |

Graphical summary of PPT, t_10-90_ and loading rate is presented in **Fig. 3**.

**Figure 3.**
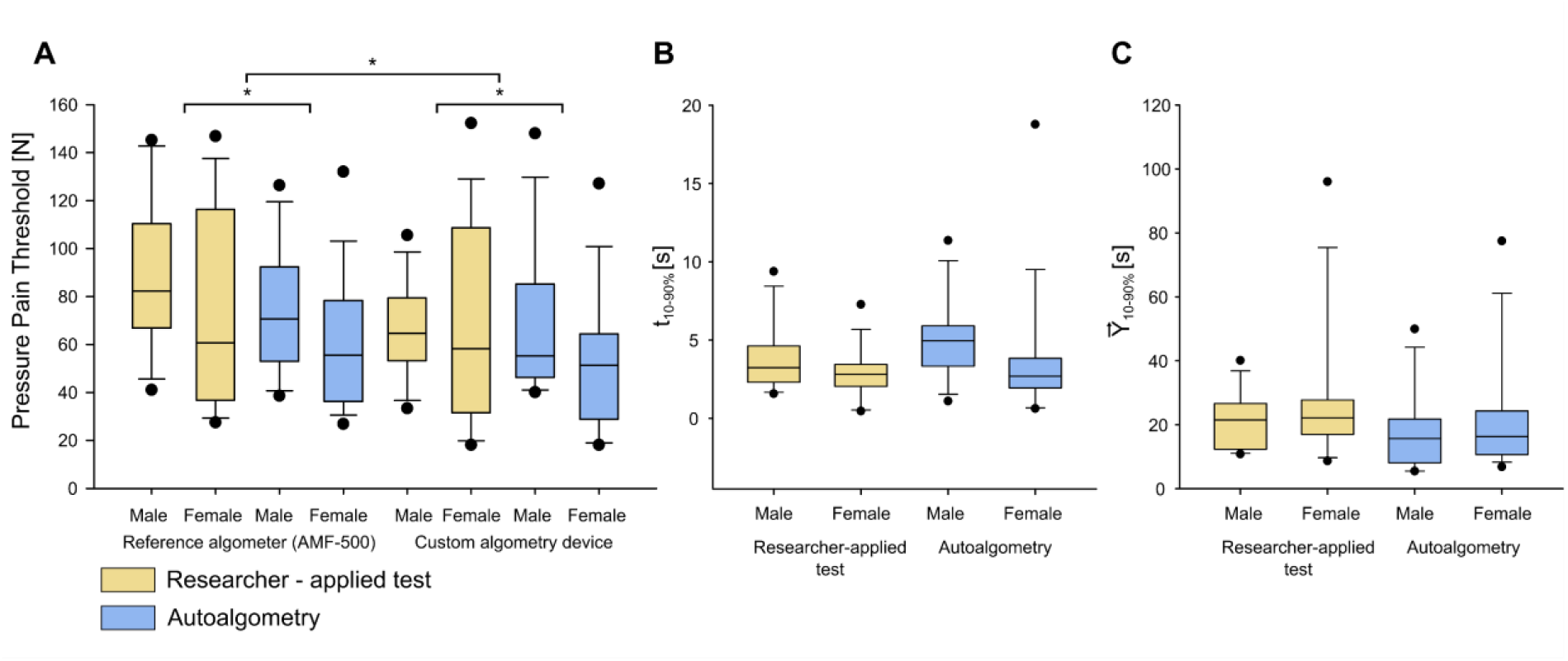
Summary of data across devices, methods of device application and volunteer’s gender. **A.** PPT values for the AMF-500 and the custom algometry device when administered by a researcher (yellow) and when self-administered (blue). **B**. t_10-90_ measured in researcher-applied and autoalgometry tests, custom algometry device. **C**. Loading rate 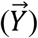 measured in researcher-applied and autoalgometry tests. ^*^ - p <0.05.

## Discussion

In this study we analyzed utilization of a previously developed pneumatic algometry device for self-administered assessment of pain-pressure threshold (PPT). Our findings suggest that this device may be suitable for use in autoalgometry. In addition, we describe considerable effects of algometry device and test methodology (researcher-administered algometry and autoalgometry) on the final PPT measurements.

The device was compared to a commercially available alternative (AMF-500 dynamometer) taken as a reference comparator in researcher-administered and self-administered tests. Pearson correlation analysis demonstrates good agreement between algometers in both tests. The two devices remain in good agreement in a Bland-Altman test. In must be emphasized that mean deference between AMF-500 obtained PPT values and custom device obtained values (all measurements conducted by researcher) is 14.3±3.8 N. This value is higher than previously reported value by Frankevich et al. (14), the possible reason for this discrepancy is that each value reported in the past was an average of 3 consecutive measurements. This difference is not expected to hamper the possibility of device implementation since absolute value of algometry is of less significance than tendency observed on measurements taken over certain time period (20).

We investigated the potential differences that may exist between measures of PPT by the comparator device (AMF-500) and the custom algometry device, as well as different methods of application. We demonstrate that PPT assessed in autoalgometry mode appears to be significantly lower for both devices and this finding is gender independent. Parenthetically, researcher which conducted the measurements was male. The effect of researcher gender is beyond the scope of this work. Of notice differences in measurements due to operator’s gender were described in literature (21). Further studies should include a male and female examiner performing tests in mixed groups to account for the possibility of examiner effect.

Lorusso et al. have previously demonstrated that the rate of pressure change has a considerable effect on the results of autoalgometry (10). To account for this, we performed evaluation of the 10-90% rise time. We demonstrate that there was no significant difference in rise time between classic algometry and autoalgometry, eliminating the possibility of application rate affecting the results to any considerable degree.

The custom algometry device validated in this work could be beneficial for regular, at-home assessment of PPT. As such, it could be adopted as a tool for monitoring progression of peripheral neuropathy. Typically, peripheral neuropathy affects sensory function first and progresses slowly over time. Severity of associated sensory alterations, numbness and neuropathic pain is time-dependent and may be used as a valuable marker of disease progression and effectiveness of therapy (22). Comprehensive neurological examination, combined with vibratory sensation testing using a tuning fork (23) and a Von Frey filament test of touch sensation (24) are typically performed in clinical setting for severity assessment. Following that, physical examination, sensory testing, laboratory tests and assessment of pain perception must be performed regularly to establish a pattern of disease progression and evaluate effectiveness of treatment (25-27).

The device described could offer a low-cost, reliable and simple way of PPT evaluation. Importantly, data obtained in such a test should not be interpreted as an alternative to conventional algometry nor should it be the sole descriptor of complex progression of this condition. Potential improvements to the proposed method include automatic increase and regulation of internal pressure combined with flexible straps or other means of fixing the device. This design would necessitate an appropriate safety mechanism such as a pressure release valve. Wireless transfer of data combined with specialized software could further enhance ease of use and help integrate PPT measurements with other markers of peripheral neuropathy (13-15) to better estimate disease progression.

This study was limited in several ways that warrant further investigation. The recruiting process resulted in a group consisting only of healthy young adults. While appropriate for a validation study, this group does not represent the population this device is intended for – namely, people affected by peripheral neuropathy. Subsequent studies should focus on patients with relevant conditions (advanced diabetes, neurodegenerative diseases, etc.) with their sensitivity, pain threshold and degree of neuropathic pain evaluated. Furthermore, algometry sites relevant to peripheral neuropathy should be tested, including plantar and palmar algometry points. Finally, to account for larger biological variation of pain-threshold in patients with neuropathic pain, a larger study group should be assembled.

## Data Availability

All data produced in the present study are available upon reasonable request to the authors

