## Supplemental tables 1 - 3 for "Application of syringe-based device for pressure pain threshold measurements under autoalgometry settings"

| **Table 1s.** Post Hoc tests for repeated measures ANOVA analysis evaluating effects of device choice and method choice on pressure-pain threshold (PPT) measurements. | | | | | | | |
| --- | --- | --- | --- | --- | --- | --- | --- |
| Comparison | |  |  | |  |  |  |
| Device | Device | Mean Difference | SE | df | | t | P_Tukey_ |
| AMF500 | Custom | 10.5 | 2.34 | 26 | | 4.50 | **< 0.001** |
| Comparison | |  |  | |  |  |  |
| Method | Method | Mean Difference | SE | df | | t | P_Tukey_ |
| Researcher-applied | Autoalgometry | 10.9 | 3.52 | 26 | | 3.08 | **0.005** |
| Comparison | |  |  | |  |  |  |
| Device × Method | Device × Method | Mean Difference | SE | df | | t | P_Tukey_ |
| AMF500, Researcher-applied | AMF500, Autoalgometry | 15.529 | 4.19 | 26 | | 3.7091 | **0.005** |
| AMF500, Researcher-applied | Custom, Researcher-applied | 15.182 | 3.68 | 26 | | 4.1217 | **0.002** |
| AMF500, Researcher-applied | Custom, Autoalgometry | 21.369 | 4.79 | 26 | | 4.4565 | **< 0.001** |
| AMF500, Autoalgometry | Custom, Researcher-applied | -0.347 | 3.57 | 26 | | -0.0972 | 1.000 |
| AMF500, Autoalgometry | Custom, Autoalgometry | 5.840 | 3.34 | 26 | | 1.7469 | 0.321 |
| Custom, Researcher-applied | Custom, Autoalgometry | 6.187 | 4.59 | 26 | | 1.3476 | 0.542 |

| **Table 2s.** Post Hoc tests for repeated measures ANOVA analysis evaluating effects of method choice and volunteer gender on T_10-90%_. | | | | | | | |
| --- | --- | --- | --- | --- | --- | --- | --- |
| Comparison | |  |  | |  |  |  |
| Method | Method | Mean Difference | SE | df | | t | P_Tukey_ |
| Researcher-applied | Autoalgometry | -1.01 | 0.550 | 26 | | -1.83 | 0.078 |
| Comparison | |  |  | |  |  |  |
| Gender | Gender | Mean Difference | SE | df | | t | P_Tukey_ |
| Male | Female | 1.14 | 0.948 | 26 | | 1.20 | 0.240 |
| Comparison | |  |  | |  |  |  |
| Method × Gender | Method × Gender | Mean Difference | SE | df | | t | P_Tukey_ |
| Researcher-applied, Male | Researcher-applied, Female | 0.934 | 0.715 | 26 | | 1.306 | 0.567 |
| Researcher-applied, Male | Autoalgometry, Male | -1.213 | 0.831 | 26 | | -1.460 | 0.475 |
| Researcher-applied, Male | Autoalgometry, Female | 0.131 | 1.050 | 26 | | 0.124 | 0.999 |
| Researcher-applied, Female | Autoalgometry, Male | -2.148 | 1.140 | 26 | | -1.884 | 0.259 |
| Researcher-applied, Female | Autoalgometry, Female | -0.804 | 0.720 | 26 | | -1.117 | 0.683 |
| Autoalgometry, Male | Autoalgometry, Female | 1.344 | 1.375 | 26 | | 0.978 | 0.763 |

| **Table 3s.** Post Hoc tests for repeated measures ANOVA analysis evaluating effects of method choice and volunteer gender on loading rate. | | | | | | | |
| --- | --- | --- | --- | --- | --- | --- | --- |
| Comparison | |  |  | |  |  |  |
| Method | Method | Mean Difference | SE | df | | t | P_Tukey_ |
| Researcher-applied | Autoalgometry | 4.41 | 1.81 | 26 | | 2.43 | **0.022** |
